# Avacopan for the Treatment of ANCA-Associated Vasculitis: The Primary Endpoints Readjudication

**DOI:** 10.64898/2026.08.13.26360315

**Authors:** David Jayne, Peter A. Merkel, Xinyu Tang, Zachary S. Wallace, Casey P. Norris, Nikieia Hayden, Sumita Bhatta, Renato D. Lopes, Amy Stallings

## Abstract

**Background:** The phase 3 ADVOCATE trial evaluated the efficacy and safety of avacopan in patients with granulomatosis with polyangiitis (GPA) or microscopic polyangiitis (MPA). Concerns raised regarding the 2019 primary endpoint adjudication process prompted a blinded, independent readjudication of all participants’ primary outcomes, the results of which are described here.

**Methods:** Patients with GPA or MPA were randomized 1:1 to receive oral avacopan 30 mg twice daily or oral prednisone on a scheduled taper, each in combination with rituximab- or cyclophosphamide-based standard of care. In 2026, the Duke Clinical Research Institute Clinical Events Classification group conducted an independent, blinded committee readjudicated the Birmingham Vasculitis Activity Score (BVAS), relapse, and remission from weeks 26 through 52 using procedures aligned with the original adjudication charter. The primary endpoints were remission at week 26 and sustained remission at week 52. As per the original analysis plan, noninferiority and superiority were declared if the lower bounds of the 95% confidence interval (CI) for the difference in the primary outcome rates between avacopan and a prednisone taper were greater than −20.0 and 0.0 percentage points, respectively.

**Results:** Among 330 participants in the intent-to-treat population, remission at week 26 was achieved by 68.1% (113/166) and 67.1% (110/164) of participants in the avacopan and prednisone taper groups, respectively, in the 2026 readjudication (adjusted difference: 2.2%; 95% CI, −7.5, 11.9), compared with 72.3% (120/166) and 70.1% (115/164) in the 2019 primary outcome adjudication (adjusted difference: 3.4%; 95% CI, −6.0, 12.8). Sustained remission at week 52 was achieved by 61.4% (102/166) and 52.4% (86/164) of participants, respectively, in the 2026 readjudication (adjusted difference: 9.8%; 95% CI, −0.3, 19.9), compared with 65.7% (109/166) and 54.9% (90/164) in the 2019 readjudication (adjusted difference: 12.5%; 95% CI, 2.6, 22.3). Concordance between the 2019 and 2026 adjudications was 95.2% for remission and 93.6% for sustained remission.

**Conclusion:** The re-analysis of ADVOCATE based on the 2026 readjudication further supports the efficacy of avacopan for GPA/MPA. Non-inferiority of avacopan versus a prednisone taper was confirmed at weeks 26 and 52 despite a median 81% reduction in glucocorticoid exposure observed in the avacopan versus prednisone taper groups. While a consistent numerical difference favoring avacopan at week 52 was observed in the 2019 and 2026 analyses, this difference did not reach statistical superiority.

## Background

Granulomatosis with polyangiitis (GPA) and microscopic polyangiitis (MPA), the two most common forms of ANCA-associated vasculitis, are rare systemic autoimmune diseases characterized by necrotizing inflammation of small-to-medium-sized blood vessels.^1,2^ Patients with GPA or MPA can experience life- and organ-threatening manifestations, particularly in the kidney or lungs, and have limited treatment options.^3,4^

Avacopan is an orally administered, small-molecule C5aR1 antagonist that was approved by the US Food and Drug Administration (FDA) in 2021 as an adjunctive treatment for adults with severe active GPA and MPA in combination with standard therapy, including glucocorticoids, on the basis of the totality of the evidence, including results of the phase 3 ADVOCATE trial.^5–8^ In ADVOCATE, participants randomized to receive avacopan achieved high rates of remission and sustained remission despite a median 81% (mean 56%) reduction in glucocorticoid exposure and lower glucocorticoid toxicity than those randomized to a standard prednisone taper.

In 2026, concerns were expressed regarding the adjudication process used in the original ADVOCATE trial, which led to retraction of the original ADVOCATE publication.^8,9^ Specifically, though the 2019 readjudication of the primary endpoints for 9 of 331 participants was conducted by a blinded adjudicator after an initial analysis of the primary outcome, the request to conduct these reassessments was prompted by an unblinded individual to ensure that the criteria defining remission were consistently applied. Following this 2019 readjudication, avacopan demonstrated non-inferiority at week 26 and superiority for achieving sustained remission at week 52 when compared with a prednisone taper; in the prior analysis of the 2019 data before unblinding and readjudication of the outcomes of 9 participants, avacopan was non-inferior to a prednisone taper at both week 26 for remission and week 52 for sustained remission but not superior at week 52.

In response to these concerns, a blinded, independent readjudication of the primary endpoints from the ADVOCATE phase 3 trial dataset was conducted in 2026 based on the original adjudication charter. Importantly, there have been no concerns raised regarding the underlying ADVOCATE data which were used to assess secondary outcomes, including kidney outcomes, quality of life, and glucocorticoid toxicity. Presented here are the results of a full re-analysis of the primary endpoints based on the readjudication.

## Methods

### Study Design

The trial design has been described previously.^10^ Briefly, ADVOCATE was a multicenter, randomized, double-blind, double-dummy, active-controlled trial conducted at 143 centers (ClinicalTrials.gov number: NCT02994927). Participants were randomized 1:1 to receive either avacopan-matching placebo and a prednisone taper (prednisone taper arm) or avacopan 30 mg twice daily and prednisone-matching placebo (avacopan arm). Both arms received background induction therapy with cyclophosphamide (followed by azathioprine or mycophenolate mofetil) or rituximab (without subsequent maintenance treatment), assigned prior to randomization at the discretion of the investigator.

The trial was performed in accordance with the principles of the Declaration of Helsinki and Good Clinical Practice guidelines. Ethics committees and institutional review boards at participating sites approved the research protocol. All participants, or a parent or guardian, provided written informed consent before enrollment. ChemoCentryx sponsored the trial and provided the study drug. Avacopan was approved by the US Food and Drug Administration in October 2021. Amgen, which acquired ChemoCentryx in October 2022, sponsored the independent 2026 readjudication.

### Primary Outcome Assessment

The primary efficacy endpoints were disease remission at week 26 (defined as a Birmingham Vasculitis Activity Score (BVAS) of 0 and no glucocorticoids for treatment of AAV within 4 weeks prior to week 26) and sustained disease remission at week 52 (defined as BVAS of 0 at week 26 and at week 52, no glucocorticoids for the treatment of AAV within 4 weeks prior to week 26 and 52, and without relapse between week 26 and 52). Participants could be deemed to have achieved remission if they had a BVAS of 0 but were using prednisone to treat adrenal insufficiency (i.e., up to 10mg/d of prednisone), asthma, an allergic reaction, or other reasons unrelated to AAV. Participants were not considered to be in remission if they had a BVAS >0 during the 4 weeks prior to week 26 (if collected for an unscheduled assessment). Relapse was defined as worsening of disease after previously achieving remission at week 26 that involved (1) one or more major items in the BVAS, or (2) three or more minor BVAS items, or (3) one or two minor BVAS items recorded at two consecutive study visits.

### Adjudication Committee

In the ADVOCATE trial, remission, sustained remission, and relapse were assessed by an independent adjudication committee (the 2019 adjudication), whose members were blinded to the treatment assignment, on the basis of the data collected throughout the trial. In 2026, a retrospective, blinded, independent readjudication (the 2026 readjudication) of the primary endpoints was conducted by the Duke Clinical Research Institute (DCRI) Clinical Events Classification (CEC) group.

The 2026 readjudication committee was instructed to adjudicate cases in accordance with an Adjudication Charter (*see Supplementary Material*) that followed processes and assessment parameters consistent with those of the original ADVOCATE Adjudication Committee Charter. The scope of the readjudication included BVAS, vasculitis damage index (VDI), relapse, and remission at weeks 26 through sustained remission at week 52, and Unscheduled and Early Termination (ET) visits occurring during this period. Amgen commissioned the 2026 readjudication but had no role in the group’s review of data or its determination of remission status.

DCRI CEC is an independent academic clinical events adjudication group with extensive experience in endpoint adjudication across all phases of clinical trials and a broad range of therapeutic areas. Physicians serving on the 2026 readjudication committee were selected based on clinical expertise in rheumatology, immunology, nephrology, and cardiology, and knowledge of clinical trial methodology and CEC processes. Physicians who were part of the original ADVOCATE adjudication committee were excluded from serving on the committee. All adjudicators were trained on the protocol, event definitions, existing source documentation, adjudication forms, and operational processes. All adjudicators were trained and certified in BVAS assessment criteria by Oxford University. Adjudications were performed using secure systems with established quality control procedures to support the generation of high-quality clinical endpoint data.

The 2026 readjudication committee used case packages with identical data as those provided to the original (2019) adjudication committee with no modification to the source data. The 2026 readjudication committee members did not have any knowledge of patient-level adjudication decisions made by the original adjudication committee. An attestation was obtained from the adjudicators confirming that they were not inadvertently unblinded to the subject identifiers or 2019 results of the nine participants based on information that was made public.

### Statistical Analysis

A supplemental statistical analysis plan was developed mirroring that of the final 2019 statistical analysis plan. The primary efficacy analyses were conducted in the intention-to-treat population, defined as all randomized participants who received at least one dose of trial medication. For each primary endpoint, the difference in the proportion of participants achieving remission was estimated using inverse-variance stratum weights^11^ and the confidence interval (CI) for the difference was estimated using the Miettinen–Nurminen method. Participants with missing data at weeks 26 or 52 were considered not to have achieved remission at the corresponding time point. For each primary endpoint, avacopan was considered noninferior to prednisone taper if the lower bound of the two-sided 95% CI for the difference (avacopan minus prednisone taper) was greater than −20 percentage points and the proportion of participants achieving remission in the prednisone taper group was at least 40 percentage points; avacopan was considered superior to prednisone taper if the lower bound of the 95% CI was greater than 0.0 percentage points. To control the overall type I error rate, the two primary endpoints were tested sequentially in the following hierarchical order: noninferiority at week 26, noninferiority at week 52, superiority at week 52, and superiority at week 26. Concordance was assessed by evaluating the percent agreement between the 2019 and 2026 adjudications.

## Results

A total of 330 participants were included in the analysis. The baseline demographics and disease characteristics of participants included in the ADVOCATE trial were well-balanced between the two treatment groups (**Table 1**).

**Table 1.** Baseline demographics and clinical characteristics*.

| Characteristic | Avacopan (N=166) | Prednisone (N=164) |
| --- | --- | --- |
| Age — yr | 61.2±14.6 | 60.5±14.5 |
| Sex — no. (%) |  |  |
| Male | 98 (59.0) | 88 (53.7) |
| Female | 68 (41.0) | 76 (46.3) |
| Race — no. (%)<sup>†</sup> |  |  |
| White | 138 (83.1) | 140 (85.4) |
| Asian | 17 (10.2) | 15 (9.1) |
| Black | 3 (1.8) | 2 (1.2) |
| Other | 8 (4.8) | 7 (4.3) |
| Body-mass index<sup>‡</sup> | 26.7 ± 6.0 | 26.8 ± 5.2 |
| Median duration of ANCA-associated vasculitis (range) — mo | 0.23 (0–362.3) | 0.25 (0–212.5) |
| Vasculitis disease status — no. (%) |  |  |
| Newly diagnosed | 115 (69.3) | 114 (69.5) |
| Relapsed | 51 (30.7) | 50 (30.5) |
| ANCA status — no. (%) |  |  |
| Antiproteinase 3 positive | 72 (43.4) | 70 (42.7) |
| Antimyeloperoxidase positive | 94 (56.6) | 94 (57.3) |
| Type of vasculitis — no. (%) |  |  |
| Granulomatosis with polyangiitis | 91 (54.8) | 90 (54.9) |
| Microscopic polyangiitis | 75 (45.2) | 74 (45.1) |
| Birmingham Vasculitis Activity Score<sup>§</sup> | 16.3±5.9 | 16.2±5.7 |
| Vasculitis Damage Index<sup>¶</sup> | 0.7±1.5 | 0.7±1.4 |
| Immunosuppressant induction treatment — no. (%) |  |  |
| Intravenous rituximab | 107 (64.5) | 107 (65.2) |
| Intravenous cyclophosphamide | 51 (30.7) | 51 (31.1) |
| Oral cyclophosphamide | 8 (4.8) | 6 (3.7) |
| Organ involvement — no. (%) |  |  |
| Renal | 134 (80.7) | 134 (81.7) |
| General | 111 (66.9) | 114 (69.5) |
| Ear, nose, and throat | 75 (45.2) | 69 (42.1) |
| Chest | 71 (42.8) | 71 (43.3) |
| Nervous system | 38 (22.9) | 31 (18.9) |
| Mucous membranes or eyes | 26 (15.7) | 40 (24.4) |
| Cutaneous | 24 (14.5) | 23 (14.0) |
| Cardiovascular | 6 (3.6) | 3 (1.8) |
| Abdominal | 4 (2.4) | 1 (0.6) |
| Glucocorticoid use during screening period |  |  |
| Use of any glucocorticoids — no. (%) | 125 (75.3) | 135 (82.3) |
| Intravenous use | 63 (38.0) | 73 (44.5) |
| Oral use | 99 (59.6) | 113 (68.9) |
| Total prednisone-equivalent dose — mg<sup>**</sup> | 907.3±1145.9 | 978.0±1157.5 |
| Daily prednisone-equivalent dose — mg<sup>**</sup> | 64.8±81.9 | 69.9±82.7 |
| * Data are shown for the modified intention-to-treat population. Plus-minus values are means ±SD. Percentages may not total 100 because of rounding. ANCA denotes antineutrophil cytoplasmic antibody.<br>† Race was reported by the patients.<br>‡ The body-mass index is the weight in kilograms divided by the square of the height in meters.<br>§ The Birmingham Vasculitis Activity Score is a composite measure of signs and symptoms in nine organ systems. Scores range from 0 to 63, with higher scores indicating more extensive disease activity.<br>¶ The Vasculitis Damage Index pertains to 11 organ systems. Values range from 0 to 64, with higher scores indicating more extensive organ damage. Patients with a new diagnosis typically have a score of 0.<br>‡ Organ involvement was based on the Birmingham Vasculitis Activity Score.<br>** The prednisone-equivalent dose includes both intravenous and oral use of glucocorticoids.<br>Source: Jayne DRW, et al. N Engl J Med. 2021;384(7):599-609; DOI: 10.1056/NEJMoa2023386.<br>The baseline characteristics presented in this table are from the original article, which has been retracted by the publisher. |  |  |

The primary outcomes, as determined by the 2019 adjudication and the 2026 readjudication processes, are shown in **Figure 1**. Based on the 2026 readjudication, remission at week 26 was achieved by 68.1% (113/166) and 67.1% (110/164) of participants in the avacopan and prednisone taper groups, respectively (adjusted difference: 2.2%; 95% CI, −7.5, 11.9; P<0.0001 for noninferiority; P=0.3301 for superiority). Using the assessments conducted by the 2019 adjudication committee, the corresponding rates of remission achieved at week 26 were 72.3% (120/166) and 70.1% (115/164), respectively (adjusted difference: 3.4%; 95% CI, −6.0, 12.8; P<0.0001 for noninferiority; P=0.2387 for superiority).

**Figure 1.**
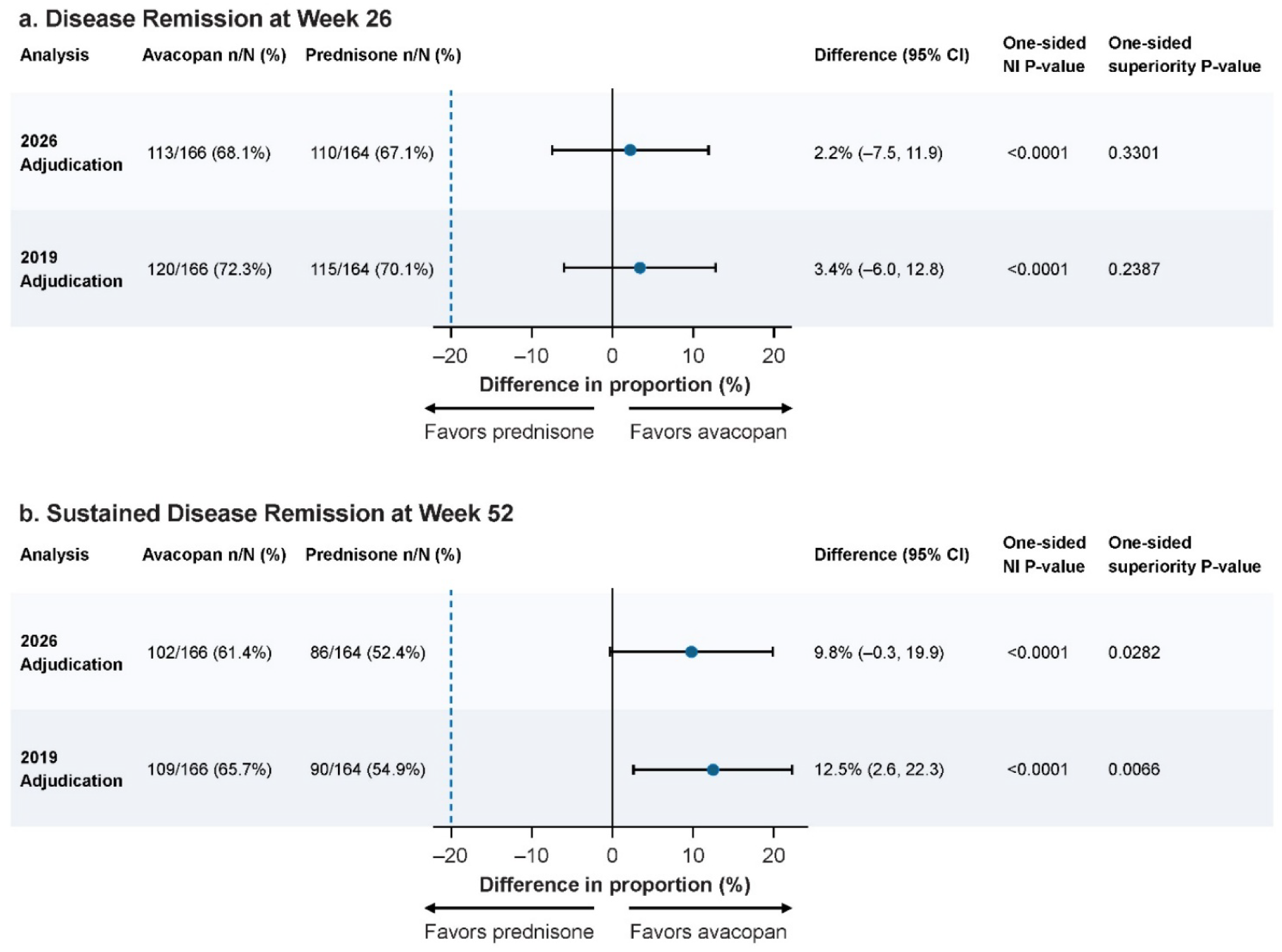
Primary Endpoints: Remission at Week 26 (a) and Sustained Remission at Week 52 (b) The 2026 adjudication refers to the retrospective, blinded, independent readjudication of the primary endpoints conducted by the Duke Clinical Research Institute Clinical Events Classification group. The 2019 adjudication refers to the results published in 2021 in the *New England Journal of Medicine* (now retracted; Jayne DRW, et al. N Engl J Med. 2021;384(7):599-609; Retraction: E. J. Rubin. N Engl J Med 2026) Confidence intervals for treatment proportions are calculated using the Clopper and Pearson Method. Two-sided 95% confidence intervals are calculated for the difference in proportions (Avacopan minus Prednisone) adjusted for randomization strata (newly diagnosed or relapsed AAV, anti-PR3 or anti-MPO ANCA, and IV rituximab or cyclophosphamide (IV or oral) standard of care treatment) using the stratified summary score test and estimate for the common difference in proportions. Non-inferiority and superiority p-values are one-sided. AAV, ANCA-associated vasculitis; ANCA, anti-neutrophil cytoplasmic antibody; IV, intravenous; MPO, myeloperoxidase; NI, non-inferiority; PR3, proteinase-3.

Using assessments from the 2026 readjudication, sustained remission at week 52 was achieved by 61.4% (102/166) and 52.4% (86/164) participants in the avacopan and prednisone taper groups, respectively (adjusted difference: 9.8%; 95% CI, −0.3, 19.9; P<0.0001 for noninferiority; P=0.0282 for superiority). In the 2019 adjudication, corresponding rates of sustained remission achieved at week 52 were 65.7% (109/166) and 54.9% (90/164), respectively (adjusted difference: 12.5%; 95% CI, 2.6, 22.3; P<0.0001 for noninferiority, P=0.0066 for superiority).

Over 52 weeks, participants receiving avacopan had a mean cumulative oral and intravenous glucocorticoid exposure of 1,676 mg prednisone equivalent, corresponding to a mean of 5 mg/day, compared with 3,847 mg (mean 13 mg/day) among participants receiving prednisone. Thus, there was a median 81% (mean 56%) reduction in glucocorticoid exposure in the avacopan versus prednisone taper arms.

Concordance between the 2019 adjudication and the 2026 adjudication analyses for the assessment of remission (at week 26) and sustained remission (at week 52) was 95.2% and 93.6%, respectively.

## Discussion

This reanalysis of the results of the ADVOCATE trial based on the 2026 readjudication of the primary endpoints demonstrated that avacopan, when used in combination with rituximab or cyclophosphamide for induction of remission, was associated with high rates of remission at week 26 and sustained remission at week 52, while substantially reducing glucocorticoid exposure in patients with GPA and MPA. There was a high degree of concordance regarding the primary endpoints assessments between the 2019 and 2026 adjudication committees, supporting the consistency of the benefits of avacopan.

As in the 2019 analysis, the lower bound of the confidence intervals in the 2026 re-analysis was well above the pre-specified noninferiority margin of −20%, providing high confidence in the noninferiority results at weeks 26 and 52. Although statistical significance for superiority of sustained remission at week 52 was not achieved with avacopan in the 2026 readjudication, the between-group difference was 9.8% (95% CI, −0.3, 19.9; P=0.0282). This consistent and strong direction was aligned with that observed in the original adjudication, the results of which were previously published (12.5%; 95% CI, 2.6, 22.3; P=0.0066). It is important to note that the ADVOCATE trial was powered to detect an 18-percentage point treatment difference and, therefore, was underpowered to demonstrate statistical significance for the smaller treatment difference of 9.8-percentage points observed in the 2026 re-analysis.

Blinded endpoint adjudication committees are commonly used in clinical trials to improve the consistency and objectivity of endpoint assessment, particularly when endpoint determination requires clinical judgment.^12^ As in ADVOCATE, adjudication committees apply prespecified criteria in accordance with an adjudication charter, and predefined procedures exist to resolve differences in interpretation among reviewers, especially in medically complex conditions such as AAV where it is necessary to determine whether a reported sign or symptoms reflects active AAV, damage, or another condition. Consequently, some differences in assessments between independent adjudication committees might occur. Unlike the 2019 adjudication committee, the 2026 adjudication committee was unable to query sites for data clarifications if members had questions regarding investigator assessments; to mitigate this limitation, the 2026 adjudication committee was provided with the query log from the 2019 adjudication committee. Despite the retrospective nature of the 2026 readjudication, concordance between the original and 2026 adjudications was high, supporting the reliability of the findings.

There remains a high unmet need for therapeutic options for treating GPA and MPA. Despite advances over the last several decades, patients with GPA and MPA continue to experience suboptimal outcomes, including relapses, chronic kidney disease, treatment-related toxicities, including those from the use of glucocorticoids and long-term immunosuppression, reduced quality of life and markedly increased mortality risk.^4^ Glucocorticoids have been a cornerstone of treatment of GPA and MPA, but cumulative glucocorticoid exposure is associated with substantial toxicities that contribute to much of the morbidity that patients with GPA and MPA experience.^13,14^

In this context, the findings from the 2019 and 2026 analyses of the ADVOCATE trial primary endpoints support the important role that avacopan can serve for patients with GPA and MPA by helping them achieve high rates of remission and sustained remission despite minimized glucocorticoid exposure. Details regarding the primary and secondary outcomes, safety, and the variance between the 2019 and 2026 adjudications will be the subject of a separate subsequent report.

In conclusion, analysis based on the 2026 readjudication of the primary endpoints of the ADVOCATE trial reaffirms the noninferiority of avacopan, as compared with a prednisone taper, for remission at week 26 and a consistent treatment effect favoring avacopan for sustained remission at week 52. These findings, combined with real world evidence^15,16^ and key secondary ADVOCATE outcomes regarding kidney outcomes,^17^ health-related quality of life^18^ and glucocorticoid-related toxicity,^19^ each of which were not impacted by the readjudication process, indicate that avacopan is beneficial to patients with GPA/MPA and remains an important therapeutic option for patients.

## ACKNOWLEDGEMENTS

None.

## COMPETING INTERESTS

**David Jayne** reports receiving honoraria or consulting fees from Alentis, Amgen, Astra-Zeneca, Autolus, BIOGEN, Boehringer, ClimbBio, CSL Vifor, Fate, GSK, Jade, Kissei, Novartis, Otsuka, and UCB. **Peter A. Merkel** reports receiving funds for the following activities in the past 2 years: Consulting: AbbVie, Alexion, Amgen, ArGenx, AstraZeneca, Boehringer-Ingelheim, Bristol-Myers Squibb, GlaxoSmithKline, Lifordi, Lilly, Melodia, Merge, Mirador, Neutrolis, Novartis, NS Pharma, Otsuka, Protallix, Q32, Quell, Regeneron, Sanofi, Sparrow, Zura. Research Support: AbbVie, Amgen, AstraZeneca, Boehringer-Ingelheim, Bristol-Myers Squibb, Electra, GlaxoSmithKline, Neutrolis, Q32, and Takeda. Stock options: Q32, Lifordi, Neutrolis, Serpass, and Sparrow. Royalties: UpToDate. **Renato D. Lopes** reports research grants or contracts from Bristol Myers Squibb, GlaxoSmithKline, Pfizer, Novartis, Ionis Pharmaceuticals Inc, Regeneron Pharmaceuticals Inc, Alnylam, Roche, Bayer, VeraDermics, Priovant Therapeutics Basilea Pharmaceutica, Theravance Biopharma Inc, Cytokinetics Inc. Funding for educational activities or lectures from Pfizer, Novartis, Novo Nordisk, and Bristol Myers Squibb. Funding for consulting from Bayer, Boehringer Ingelheim, Bristol Myers Squibb, Novo Nordisk, Novartis, and Pfizer. **Xinyu Tang, Zachary S. Wallace and Sumi Bhatta** are employees of, and hold stock in, Amgen, Inc. **Amy Stallings, Casey P. Norris, Nikieia Hayden** do not have any conflict of interests.

## FUNDING

This work was funded by Amgen, Inc.

## DATA AVAILABILITY

The data underlying the analyses described in this report are not publicly available. Following completion of the applicable review and governance processes, qualified researchers may submit requests for access to study data for scientifically sound research purposes. Requests will be evaluated by an internal data review committee according to predefined scientific, ethical, legal, and contractual criteria. Access, if approved, will be provided under an appropriate data-sharing agreement and in accordance with applicable laws, regulations, and participant privacy protections.

## AUTHOR CONTRIBUTIONS

DJ and PAM conceived the original design of the ADVOCATE trial. SB conceived of the design of the re-adjudication of the ADVOCATE primary outcomes. AS, XT, CPN, RDL, NH, and SB contributed to the methodology. AS, CPN, RDL, and NH contributed to investigation. XT contributed to software, formal analysis, and data visualization. AS, CPN, RDL, and NH contributed to data curation. XT and ZSW drafted the original manuscript. DJ, PAM, AS, ZSW, CPN, RDL, NH, and SB reviewed and edited the draft and provided critical feedback. AS, CPN, RDL, and NH provided supervision, and NH project administration support.

## Notes

### Clinical Trial

NCT02994927

### Author Declarations

The following IRBs provided ethics approval to the original phase 3 trial: 1. Advarra Institutional Review Board: 6940 Columbia Gateway Drive, Suite 110 Columbia, MD 21046, USA 2. Partners Human Research Committee 399 Revolution Drive, Suite 710 Somerville, MA 02145, USA 3. Schulman Institutional Review Board, 4445 Lake Forest Drive, Suite 300 Cincinnati, OH 45242, USA 4. Brany Institutional Review Board: 1981 Marcus Avenue, Suite 210 Lake Success, NY 11042, USA 5. Western Institutional Review Board: 1019 39th Avenue SE Suite 120, Puyallup, WA 98374, USA 6. University of Kansas Medical Center: 3901 Rainbow Blvd. Kansas City, KS 66160, USA 7. University of Chicago Institutional Review Board: 5841 S. Maryland Ave. MC7132, I-625 Chicago, IL: 60637, USA "8. Columbia University Medical Center Institutional Review Board 154 Haven Ave., 1st Floor New York, NY 10032, USA" 9. University of California San Francisco Committee on Human Research: 3333 California Street, Suite 315 San Francisco, CA 94118, USA 10. Lifespan Research Protection Office: CORO West, Suite 1300 One Hoppin Street Providence, Rhode Island 02903, USA 11. Mayo Clinic Institutional Review Board: 200 First Street SW Rochester, MN 55905, USA "12. Georgetown-Howard Universities Center for Clinical and Translational Science Institutional Review Board 3900 Reservoir Road NW, SW 104 Med Dent Building Washington, DC 20007, USA" 13. Human Research Protection Office: Washington University in St. Louis, 660 S. Euclid Ave., Campus Box #8089, St. Louis, MO 63110, USA 14. Chesapeake Research Review, Inc.: 6940, Columbia Gateway Drive, Suite 110 Columbia, MD 20146-3403, USA 15. NYU Winthrop Hospital, Institutional Review Board: 222 Station Plaza North Suite 521 Mineola, NY 11501, USA 16. Cedars-Sinai Medical Center Institutional Review Board: 6500 Wilshire Blvd. Suite 1800 Beverly Hills, CA 90048, USA 17. Office of the Human Research Protection Program: 10889 Wilshire Blvd, Suite 820, Los Angeles, CA 90095-1406, USA 18. John Hopkins Medicine Institutional Review Board: 1620 McElderry Street Reed Hall B-130 Baltimore, Maryland 21205-1911, USA 19. The Cleveland Clinic Foundation Institutional Review Board: 9500 Euclid Ave. OS-1 Cleveland, OH 44195, USA 20. University of Utah Institutional Review Board: 75 South 2000 East, Salt Lake City, Utah 84112, USA 21. Boston University Medical Campus Institutional Review Board 560 Harrison Ave, Third Floor, Suite 300 Boston, MA 02118, USA "22. East Carolina University - University & Medical Center Institutional Review Board 4N70 Brody Medical Sciences Building, MailStop 682 600 Moye Boulevard Greenville, NC 27834, USA" "23. University of Kentucky Office of Research Integrity 315 Kinkead Hall Lexington, KY 40506, USA" "24. University of Minnesota Human Research Protection Program 420 Delaware St SE, D528 Mayo Memorial Building, MMC 820 Minneapolis, MN 55455, USA" 25. Mount Sinai Ethics Review Board 700 University Avenue Suite 8-600 Toronto, ON MSG 1Z5, Canada "26. Conjoint Health Research Ethics Board: 2500 University Drive NW 3rd Floor MacKimmie Library Tower Calgary, Alberta T2N 1N4, Canada" "27. Comite d'ethique de la recherche du CIUSSS de l'Est-de-l'Ile-de-Montreal 5415 Boul de l'Assomption, Suite 4158, 4th floor, Pavillon Rachel-Tourigny, Montreal., Quebec HlT2M4, Canada" "28. CIUSSS de l'Est-de-l'ile-de-Montreal - installation Hopital Maisonneuve-Rosemont 5415 boul. l'Assomption, Montreal, Quebec HlT 2M4, Canada" 29. UBC PHC REB: 10th Floor - 1190 Hornby Street, Vancouver, BC V67 2K5 "30. Hamilton Integrated Research Ethics Board 293 Wellington Street North Hamilton, Ontario L8L8E7, Canada" "31. Le Comite d'ethique de la recherche du crusss de l'Estrie - CHUS: 3001, 12e avenue Nord Sherbrooke, Quebec J1H5N4, Canada" "32. Comite d'ethique de la recherche du CIUSSS de l'Est-de-l'ile-de Montreal: 66, Sainte-Catherine Est Montreal, Quebec H2X IK6, Canada" "33. Comite Ethique de la recherche du CIUSSS de l'Est-de-l'Ile -de-Montreal 5415,Boulevard de l'Assomption Montreal, Quebec H1T2M4, Canada" 34. Ethikkommission der Medizinischen Universitat Graz: LKH-Universitatsklinikum, Auenbruggerplatz 2 Graz, A-8036, Austria "35. Comite d'Ethique Hospitalo-Facultaire Saint-Luc: Avenue Hippocrate 55.14, Tour Harvey - Niveau 0 Bruxelles 1200, Belgium" "36. Comite d'Ethique Hospitalo-Facultaire Saint-Luc: Promenade de l' Alma 51 Bte Bl.43.03 1200 Bruxelles, Belgium" 37. CEC: Eticka komise Fakultni nemocnice Kralovske Vinohrady, Srobarova 1150/50, Praha 10 100 34, Czech Republic 38. De Videnskabsetiske Komiteer for Region Hovedstaden, Kongens Vaenge 2 Hillerod,3400, Denmark 39. Comite de Protection des Personnes Ile-De-France IV: Hopital Saint Louis - Porte 5 du Carre Historique, 1 avenue Claude Vellefaux, Paris 75475 cedex 10, France 40. Ethikkommission der Medizinischen Fakultat Heidelberg: Alte GlockengieBerei 11/1, Heidelberg BW 69115, Germany 41. Research Ethics Committee SJH/AMNCH Tallaght Hospital, Dublin 24, Ireland 42. CEC - Comitato Etico Regionale della Liguria - Sezione 3, Largo Rosanna Benzi 10 Genova, GE 16132, IT 43. Comitato Etico Regionale della Liguria - Sezione 3, IRCCS AOU San Martino 1ST Largo Rosanna Benzi 10 Genova, NAP 16132, Italy 44. Comitato Etico Regionale della Liguria - Sezione 3, Largo Rosanna Benzi 10, Genova GE 16132, Italy 45. METc UMCG: Hanzeplein 1, Groningen, Groningen 9713 GZ, The Netherlands "46. Regional komite for medisinsk og helsefaglig forskningsetikk (REK) Sor-ost: Gullhaugveien 1-3 Oslo 0484, Norway" 47. Comite Autonomico de Etica de la Investigacion de Galicia (CAEI de Galicia): Edificio Administrativo San Lazaro, C/ San Lazaro, s/n,Santiago de Compostela, A Coruna 15703, Spain "48. Regionala Etikprovningsnamden i Stockholm Widenstromska huset, Tomtebodavagen 18A Solna, 171 65, Sweden" 49. East of England - Cambridge South Research Ethics Committee: The Old Chapel, Royal Standard Place Nottingham, NG1 6FS, UK 50. East of England - Cambridge South Research Ethics Committee, The Old Chapel, Royal Standard Place, London, NG1 6PS, UK 51. Central Adelaide Local Health Network (CALHN) HREC: Roma Mitchell House, Level 3, 136 North Terrace, Adelaide, SA 5000, Australia 52. UnitingCare Health Human Research Ethics Committee: Ground Floor, Moorlands House, 451 Coronation Drive, Auchenflower, QLD 4066, Australia "53. Sir Charles Gairdner Hospital Human Research Ethics Committee: Hospital Avenue Nedlands, WA 6009, Australia" 54. Health and Disability Ethics Committees 133 Molesworth Street, PO Box 5013 Wellington 6011, New Zealand 55. Ethikkommission Ostschweiz Haus 37: St. Gallen, St. Gallen 9007, Switzerland "56. Egeszsegugyi Tudomanyos Tanacs Klinikai Farmakologiai Etikai Bizottsaga: Alkotmany u. 25. Budapest 1054 Hungary" 57. Kobe University Hospital Institutional Review Board: 7-5-2,Kusunoki-Cho,Chuo-Ku Kobe-City, Hyogo 650-0017, Japan "58. Okayama University Hospital Institutional Review Board: 2-5-1 Shikata-cho, Kita-ku Okayama-city, Okayama 700-8558, Japan" 59. National Hospital Organization Nagoya Medical Center Institutional Review Board: 4-1-1 Sannomaru Naka-ku, Nagoya-city, Aichi 460-0001, Japan "60. Juntendo University Shizuoka Hospital Institutional Review Board: 1129, Nagaoka Izunokuni-shi, Shizuoka 410-2295, Japan" "61. University of Miyazaki Hospital Institutional Review Board: 5200 Kiyotakecho Kihara Miyazaki-city, Miyazaki 889-1692 Japan" 62. Shimane University Hospital Clinical research review board: 89-1, Enya-cho Izumo, Shimane 693-8501, Japan 63. Saitama Medical University Hospital Institutional Review Board: 38, Morohongo, Irumagun Moroyamamachi Saitama 350-0495, Japan "64. Kagawa University Hospital Institutional Review Board: 1750-1, Ikenobe, Miki-cho Kita-gun, Kagawa 761-0793, Japan" 65. Hiroshima University Hospital Institutional Review Board, Kasumi 1-2-3 Minami-ku Hiroshima-city, Hiroshima 734-8551, Japan 66. Institutional Review Board of Saitama Medical Center, 1981 Kamoda Kawagoe, Saitama 350-8550, Japan 67. National Hospital Organization Osaka Minami Medical Center Institutional Review Board: 2-1 Kido Higashimachi, Kawachi Nagano-shi, Osaka 586-8521, Japan "68. Hokkaido University Hospital Institutional Review Board: Kital 4, Nishi 5; Kita-ku Sapporo, Hokkaido 060-8648, Japan" 69. Akita University Hospital Institutional Review Board: 44-2, Hasunuma; Hiroomote Akita-City, Akita 010-8543, Japan 70. Kyorin University Hospital Institutional Review Board: 6-20-2, Shinkawa Mitaka, Tokyo 181-8611, Japan "71. Osaka Medical College Hospital Institutional Review Board: 2-7 Daigaku machi Takatsuki City, Osaka 569-8686, Japan" "72. Teikyo University Hospital Istitutional Review Board: Kaga 2-11-1 Itabashi-ku, Tokyo 173-8606, Japan" 73. Yokohama City University Hospital Institutional Review Board: 3-9 Fukuura, Kanazawa-Ku Yokohama, Kanagawa 236-0004 Japan "74. Hamamatsu University Hospital Institutional Review Board: 1-20-1 Handayama, Higashi-ku Hamamatsu-City, Shizuoka 431-3192, Japan" 75. National Hospital Organization Yokohama Medical Center Institutional Review Board 3-60-2, Harajuku, Totsuka-ku, Yokohama-city, Kanagawa 245-8575, Japan 76. National Hospital Organization Chiba East Hospital Institutional Review Board: 673 Nitona-cho, Chuo-ku, Chiba-shi, Chiba 260-8712, Japan "77. National Hospital Organization Tokyo Medical Center Institutional Review Board: 2-5-1,Higashigaoka Meguro-ku, Tokyo 152-8902, Japan" "78. Nagoya City University Graduate School of Medical Sciences and Nagoya City University Hospital Institutional Review Board 1-Kawasumi, Mizuho-cho, Mizuho-ku Nagoya, Aichi 4678602, Japan" 79. Keio University Hospital Institutional Review Board: 35 Shinanomachi Shinjuku-ku, Tokyo 160-8582, Japan 80. Toho University Omori Medical Center Institutional Review Board: 6-11-1, Omori-nishi, Ota-ku, Tokyo 143-8541, Japan 81. Juntendo University Hospital Institutional Review Board: 3-1-3 Hongo, Bunkyo-ku, Tokyo 113-8431, Japan 82. National Hospital Organization Kanazawa Medical Center Institutional Review Board: 1-1, Shimoishibiki-machi, Kanazawa, Ishikawa 920-8650, Japan 83. Teikyo University Chiba Medical Center Institutional Review Board: 3426-3, Anesaki, Ichihara-city, Chiba 299-0111, Japan 84. Tazuke Kofukai Medical Research Institute Kitano Hospital Institutional Review Board: 2-4-20, Ohgimachi, Kita-ku, Osaka-city, Osaka 530-8480, Japan 85. Toyama University Hospital Institutional Review Board: 2630 Sugitani, Toyama-shi, Toyama 930-0194, Japan 86. Okayama saiseikai General Hospital Institutional Review Board: 2-25, Kokutaicho, Kita-ku, Okayama-city, Okayama 700-8511, Japan 87. University of Tsukuba Hospital Institutional Review Board: 2-1-1, Amakubo, Tsukuba-city, Ibaraki 305-8576, Japan

