## Supplemental Material for "Avacopan for the Treatment of ANCA-Associated Vasculitis: The Primary Endpoints Readjudication"

### DCRI Clinical Events Classification (CEC) Charter for ADVOCATE Study

#### Protocol CL010_168

A Randomized, Double-Blind, Placebo-Controlled, Phase 3 Study to Evaluate the Safety and Efficacy of CCX168 (Avacopan) in Patients with Anti-Neutrophil Cytoplasmic Antibody (ANCA)-Associated Vasculitis Treated Concomitantly with Rituximab or Cyclophosphamide/Azathioprine

D1: D2744

Charter Effective Date:

The Effective date is the date of the last signature on the Document Approval Form

Version 2.0

[redacted]

**REQUIRED** *My signature confirms that I have reviewed, approved and will follow the CEC Charter.*

| Title | Name | Signature/Date |
|---|---|---|
| Lead CEC Coordinator | [redacted] | |
| CEC Project Leader | | |
| DCRI Project Leader | | |
| CEC Co-Principal Investigator | | |
| CEC Co-Principal Investigator | | |

**OPTIONAL** *My signature confirms that I have reviewed the CEC Charter*

| Title | Name | Signature/Date |
|---|---|---|
| DCRI Principal Investigator | N/A | |
| Clinical Operations Project Leader | N/A | |
| Data Management Representative | N/A | |
| Project Statistician | N/A | |
| Sponsor Representative | [redacted] | |
| Sponsor Representative | N/A | |
| Other: | N/A | |

#### Document Version History

| Version Number | Effective Date | Authored/ Revised By | Reason for Revision |
|---|---|---|---|
| 1.0 | 25Mar2026 | [redacted] | Initial document |
| 2.0 | Date of last signature | | • Updated Section 5.3 and Section 7 to reflect tiebreaker assessments may be conducted by two committee members.<br>• Updated Section 5.4 to clarify that QC may be performed by an external Physician Reviewer. |

[redacted]

[redacted]

#### 1. Acronyms

| Acronym | Definition |
|---|---|
| AAV | Anti-Neutrophil Cytoplasmic Antibody (ANCA)-Associated Vasculitis |
| AF | Adjudication Form |
| ANCA | Anti-Neutrophil Cytoplasmic Antibody |
| BVAS | Birmingham Vasculitis Activity Score |
| CEC | Clinical Events Classification |
| Co-PI | Co-Primary Investigator |
| CT | Computed Tomography |
| CTC | Clinical Trials Coordinator |
| CV | Curriculum Vitae |
| DCRI | Duke Clinical Research Institute |
| EDC | Electronic Data Capture |
| ET | Early Termination |
| FDF | Financial Disclosure Form |
| eGFR | Estimated Glomerular Filtration Rate |
| GN | Glomerulonephritis |
| GPA | Granulomatosis with Polyangiitis |
| MPA | Microscopic Polyangiitis |
| PHI | Protected Health Information |
| PL | Project Leader |
| QC | Quality control |
| RBC | Red Blood Cells |
| SOP | Standard Operating Procedures |
| SAP | Statistical Analysis Plan |
| SSAP | Supplemental Statistical Analysis Plan |
| VDI | Vasculitis Damage Index |
| WBC | White Blood Cells |

[redacted]

#### 2. ROLE of the DCRI CEC

The Duke Clinical Research Institute (DCRI) Clinical Events Classification (CEC) group has been requested to perform a retrospective re-adjudication in collaboration with Amgen. The DCRI CEC group creates and maintains the CEC Charter; develops and executes the CEC workflow and processes in collaboration with the Amgen.

The CEC group will perform re-adjudication of the Primary Endpoints in Protocol CL010_168 V 4.0 using definitions and rules specified in the Protocol, Statistical Analysis Plan (SAP), and Supplemental Statistical Analysis Plan (SSAP). The scope of work includes BVAS, Relapse, and Remission assessments at applicable timepoints including Weeks 26 through 52, including Unscheduled and Early Termination (ET) visits occurring during this period.

**For BVAS, the particular focus will be on the following:**

1. Whether disease remission was achieved at Week 26 (defined as a BVAS of 0 and not taking glucocorticoids for treatment of AAV within 4 weeks prior to Week 26). Note: There must be no BVAS>0 during the 4 weeks prior to Week 26 (if collected for an unscheduled assessment).

2. Whether sustained disease remission was achieved at Week 52. Sustained remission at Week 52 is defined as remission at Week 26 without relapse to Week 52 and remission at Week 52 (BVAS of 0 and not taking glucocorticoids for treatment of AAV for 4 weeks prior to Week 52).

3. Whether relapses have occurred between Week 26 and Week 52 (patients who relapse after remission has been achieved at Week 26 are considered treatment failures for the Week 52 sustained remission endpoint analysis).

For assessment of Vasculitis Damage Index (VDI), the CEC group will review VDI scores at Weeks 26 and 52 and any other applicable visits (such as early termination) and determine if the site investigator scoring is correct. The CEC reviewers will note any discrepancies in VDI scoring on the corresponding adjudication form.

The following rules will apply to the BVAS and/or VDI assessments:

- Disease duration should be calculated from date of diagnosis for all patients
- For all study visits, record the disease activity present within the 28 days prior to the visit.
- VDI is always 0 for newly diagnosed patients
- Regarding 'other items', sites will select in EDC whether the 'other item' is major or minor and indicate the applicable organ system
- Red blood cells (RBC) casts and glomerulonephritis have been permitted as an 'other item' and would score as a '6'. The score for new/worse hematuria would also be '6' (only RBC/Glomerulonephritis (GN) or hematuria should be marked, but not both).
- Other 'other items' should be scored as a '2' for minor and a '4' for major

[redacted]

Events will be adjudicated by the CEC group according to pre-specified criteria. The members of the CEC group will remain blinded to treatment assignment throughout the adjudication process. It is assumed the CEC adjudicated data will be used in the trial final analysis unless otherwise stated in the protocol, SAP, and SSAP.

Any amendments to this Charter will be approved in the same manner as the original Charter development process, and per CEC standard operating procedures (SOPs) governing this process. Any revisions deemed necessary by the CEC Co-Primary Investigators (Co-PIs), or the Sponsor will be subject to review by both parties prior to being adopted into the Charter as an amendment.

#### 3. Protocol Summary

The primary objective is to evaluate the efficacy of CCX168 (avacopan) to induce and sustain remission in patients with active anti-neutrophil cytoplasmic antibody (ANCA)-associated vasculitis (AAV), when used in combination with cyclophosphamide followed by azathioprine, or in combination with rituximab.

#### 4. Role of the CEC Team

The CEC Co-PIs will work with the CEC Project Leader (PL) to provide guidance and thought leadership throughout the life of the trial. The CEC Co-PIs will also assist with the selection of the CEC Physician Reviewers, provide project oversight, and can also participate in the adjudication of individual events.

##### 4.1 CEC Physician Reviewers

**Selection of CEC Physician Reviewers:**
The CEC Reviewers will be composed of a team of physicians who will be selected from Duke University/DCRI, or other institutions. Sponsor representatives will not serve as a CEC Reviewer, or participate in the adjudication of any events. However, the Sponsor may recommend MDs for inclusion into the Committee, at their discretion.

Accordingly, any adjudicator who participated in the initial ADVOCATE trial, cannot participate in CEC processes or case adjudication for the current Charter.

**Qualifications of the CEC Physician Reviewers:**
All CEC Physician Reviewers will be selected based on:

- Clinical expertise in one or more of the following therapeutic areas:
  - Rheumatology, immunology, nephrology, and cardiology
- Knowledge of clinical trial methodology and CEC processes
- Availability and commitment to the goals and timelines of the trial
- Independence from ADVOCATE trial conduct:
  - Study Investigator or Sub-Investigator

[redacted]

- Site Investigator or Sub-Investigator
    - DSMB Member
    - Study Medical Monitor
    - Voting members of Ethics or Steering Committees
    - Member of original ADVOCATE Adjudication Committee

The CEC Co-PIs will evaluate the candidate's credentials. After review and approval by the CEC Co-PI's, the reviewer credentials are reviewed and approved by the Sponsor. The Sponsor is responsible for assessing any potential conflicts of interest. DCRI CEC project team will have each Physician Reviewer complete a financial disclosure form (FDF) (DCRI document ID: OP-F1-014).

A list of Physician Reviewers will be maintained including the start dates and end dates of their work on the study. These documents will be kept in the study eTMF.

DCRI CEC Group will maintain a documentation file of all reviewer curriculum vitae (CV) for record keeping purposes.

All CEC Physician Reviewers will undergo training on the CL010_168 protocol, consisting of all the necessary information that the member will need to know to effectively adjudicate events for the trial, including but not limited to:

- Summary of the protocol
- Detailed list and definition of each event to be adjudicated
- Overview of the source documentation and dossier contents needed
- Adjudication Form(s) to be completed for the respective event types
- Expected timelines for review
- CEC operational processes

In addition, all DCRI Physician Reviewers must be certified on BVAS and VDI assessment criteria, proctored and measured by Oxford University. Physician Reviewers will not be able to begin case adjudication without BVAS-specific certification from the Oxford team.

The training material is produced and provided to the CEC Physician Reviewers as a collaborative effort by DCRI, Amgen, and/or their operational partners.

##### 4.2 CEC Team Members

| Role | Responsibilities |
|---|---|
| CEC Faculty Director/CEC Co-Primary Investigator | • Oversee the CEC process<br>• Provide strategic oversight as well as support to the CEC PI and the DCRI CEC project team |

[redacted]

| Role | Responsibilities |
|---|---|
| CEC Co-Primary Investigator | • Provide oversight:<br> o Development and finalization of the CEC charter<br> o Operational support<br> o Adjudication and quality control processes defined within the Charter<br> o Provide updates as needed to the trial stakeholders |
| CEC Physician Reviewer | • Adjudicate events adhering to Charter event definitions<br>• Participate in committee meetings |
| CEC Project Leader | • Manage:<br> o Timelines and processes including project deliverables milestones, and archival<br> o Scope of work<br> o Project specific communication/escalation<br> o Study staff training<br> o Ensure consistency in work<br> o Performance and process improvement strategies in conjunction with the Clinical Trial Coordinator (CTC)<br> o Authorize study team member access<br>• Ensure CEC projects are audit compliant<br>• Primary Sponsor contact in concert with the CTC on CEC related tasks and topics. (via meetings, phone calls, emails, online web-based teleconferences) |
| CEC Clinical Trial Coordinator | • Collaborate in the development of CEC processes<br>• Provide the sites with the necessary tools and training<br>• Participate in CEC related meetings or teleconferences<br>• Development and finalization of the CEC Charter and associated documents in concert with the CEC PL and CEC PI<br>• Facilitate the CEC adjudication meetings including requests for additional information<br>• Manage daily operations, training and workflow, to ensure timelines are met in collaboration with the CEC PL<br><br>*Note: The Lead CTC is responsible for the management of the CTC team if more than one CTC is on the team, or is the only CTC on a smaller project. The Lead CTC can authorize study team member access for the trial.* |

[redacted]

| Role | Responsibilities |
|---|---|
| CEC Clinical Trial Specialist | • Original adjudication packet retrieval via Sponsor-provided secure location<br>• Event tracking<br>• As needed, in conjunction with the Sponsor, perform/confirm all non-critical protected health information (PHI) is redacted from all received packets<br>• Create electronic casebook folders for archival packets |
| CEC Clinical Data Specialist | • Manage both incoming and outgoing document transfer specifications with the Sponsor and/or their DM partners<br>• Manage all technical aspects of the study and oversee technical study-related communication to appropriate stakeholders<br>• Troubleshoot, resolve and triage issues with applications and reports |

### 5. Event Adjudication

The CEC Physician Reviewers will be blinded to the original adjudication results. The DCRI CEC adjudication forms will be provided to the reviewers via Adobe Sign.

The Step-by-Step process for each patient assessment for the re-adjudication effort will be as follows:

- Step 1 – Review patient profiles including BVAS, VDI and steroid use
- Step 2 – Assess remission at Week 26
- Step 3 – Assess sustained remission at Week 52

#### 5.1 Data Supporting Adjudication

The CEC will review patient profiles that were utilized in the original adjudication performed for the CL010_168 trial. The CEC will also have access to a query and answer log generated from the original adjudication committee which will be utilized as an additional reference. Each CEC data package (also known as a Patient Profile) will contain data for one patient. Profiles generated by Medpace will contain the following data that was collected within the ADVOCATE study electronic data capture (EDC) (if available at the time that the Patient Profile was generated):

- Stratification Variables
- Baseline and Demographics Information
- Disposition
- Medical History
- Physical Exams
- Adverse Events

[redacted]

- Chest X-Ray/Chest Computed Tomography (CT)
- Prior and current concomitant Medications
- Concomitant non-study-supplied glucocorticoid medications
- BVAS and VDI (as assessed by the Investigator)
- Relapse information

##### Central Laboratory Data

In addition, the following Central Laboratory Data will be incorporated into each data package (Patient Profile), as available:

- Chemistry
    - Albumin
    - Urinary Albumin: Creatinine Ratio
    - Serum Creatinine
    - Estimated Glomerular Filtration Rate (eGFR)
- Hematology
    - Hemoglobin
    - Lymphocyte count
    - Neutrophil count
    - Platelet count
- Urinalysis
    - Blood
    - Nitrite
    - Protein
    - Urinary Red Blood Cells (RBC)
    - Urinary White Blood Cells (WBC)

#### 5.2 CEC Adjudication Primary Endpoints

CEC members will adjudicate each event using the protocol-specified endpoint criteria, based on the preponderance of the evidence and clinical knowledge and experience.

The Primary Endpoints for the re-adjudication efforts are:

1. *Disease Remission at Week 26*
    - All of the following must be met as determined by the CEC
        - BVAS = 0
        - No glucocorticoids* for treatment of AAV within 4 weeks prior to Week 26
        - No BVAS > 0 during the 4 weeks prior to Week 26 (if collected for an unscheduled assessment)
2. *Sustained Disease Remission at Week 52*
    - All of the following must be met as determined by CEC:
        - Valid remission at Week 26
        - No relapse between Weeks 26 and 52
        - BVAS = 0 at Week 52
        - No glucocorticoids* for treatment of AAV within 4 weeks prior to Week 52

[redacted]

***Glucocorticoid Rules/Conventions:**

Glucocorticoid use in the 4 weeks prior to the BVAS assessment at Weeks 26 and 52 should be considered when assessing remission at those time points.

Note that subjects are permitted to receive [1] low doses of oral glucocorticoids (no more than 10 mg per day) for treatment of adrenal insufficiency or [2] glucocorticoid treatment for other (non AAV) conditions, e.g., allergic reaction, and will be classified as responders for purposes of assessment of the primary endpoints if all other requirements for response are met.

#### 5.3 Event Documentation

The CEC Physician Reviewer or Committee will complete Adjudication Forms (AFs) for each applicable visit - Remission AF, Relapse AF, and BVAS AF - for each patient as packages are reviewed and an adjudication decision is made.
CEC AFs must have patient ID, visit identifier, and applicable footers for identification purposes which are to be completed by the CEC team.

As shown in the process workflow (Section 7.0), event adjudication for the study will follow a two-phase process: Phase I and Phase II.

Phase I adjudication is the individual review of the same event – in this case, patient timepoints - by two (2) separate adjudicators. If the 2 adjudicators agree on all key adjudication variables for an event (remission or relapse), that event is considered complete. For discordance in key variables within a remission timepoint assessment, the event will move to Phase II.

Phase II adjudication will be a “tiebreaker” assessment between two CEC Physician Reviewer Committee members. Adjudication results based on that tiebreaker assessment will be deemed the final results for that timepoint.

#### 5.4 Quality Control (QC) of Adjudication

The CEC QC plan for the ADVOCATE study will include a minimum of 10% random sample of the total number of adjudicated events. The purpose of QC is to ensure consistency within the adjudication process. The sample will be generated by the DCRI CEC Data Specialist. The adjudicated events selected in the sample are re-reviewed by a CEC Co-Chair or external Physician Reviewer, who is blinded to the original adjudicated results. The QC will be completed at 1 timepoint during the trial.

QC definition of major vs. minor discrepancy has been classified as follows:

Major Discrepancy:
- Did primary study outcome (and its components) occur? (yes vs. no)

Minor Discrepancy:
- Any disagreement not involving the primary study outcome (and its components)

[redacted]

Major discrepancies between the original adjudication decision and the QC will be reviewed by the CEC Co-PI, with a final decision made on whether to update the adjudicated event results for said event(s).

Potential actions based on the results of a QC review include:

- Continuing the CEC process without modifications
- The CEC Chair/PI or designee will re-educate the CEC Reviewers and reinforce review processes (retraining documentation maintained in the form of emails, slide deck, Reviewer manual, group training, etc.)
- Perform additional QC (e.g., increase QC sample size)

The results of the QC review are summarized in a blinded manner using the QC Project Summary Report, reviewed by the CEC Co-PIs and CEC PL, and findings sent to the Sponsor for review, comment, and eventual approval by all parties.

### 6. Study Assessments

#### 6.1 Birmingham Vasculitis Activity Score (BVAS) Version 3

The BVAS form has been completed by study site Investigators according to the schedule provided in the protocol Time and Events Table, including at Week 26 and Week 52. In Protocol Section 12.3, instructions for completion, and the BVAS organ systems and items within each system are provided. The "major" BVAS items are listed in bold italics. Calculation of the BVAS score was not performed by the Investigators, but was performed in the EDC system.

BVAS data recorded by Investigators (including Week 26 and Week 52) will be adjudicated, according to timepoints specified in this adjudication charter.

The CEC Physician Reviewer will complete an entire BVAS AF for each visit at which the Adjudicated BVAS responses differ from the Investigator's BVAS responses. Comments will be entered onto the AF as appropriate to document significant discussions and/or rationale, that should be included with the final adjudication dataset. All adjudication considerations must be meticulously documented with rationale.

On the BVAS AF:

- **Major Items** on the BVAS form (marked with "#" on the form) do **not** require completion by the Physician Reviewer.
- The question: **"Is this the patient's first assessment?"** on the BVAS AF does **not** require completion by the Physician Reviewer.
- For BVAS forms, if the Physician Reviewer agrees with the site investigator's scoring, the Physician Reviewer can select the "Agree" box and sign either the front page of the form, or on the signature line of the last page.
- Alternatively, the CEC Chair or designee may complete the CEC-specific BVAS Scoring Agreement Form and tick the applicable BVAS assessments (or enter "Early Term" as applicable). If this form is completed, the individual BVAS form does not require completion.

[redacted]

- Note if the Physician Reviewer disagrees with the site investigator scoring, “disagree” should be selected and the forms will be completed as appropriate.

The following instructions need to be followed to complete the BVAS:

- Record only symptoms/signs ascribed to the presence of active AAV (GPA or MPA)
- Distinguish between active disease captured on BVAS versus damage captured on the VDI instrument (see Section 6.4 Vasculitis Damage Index).
- “Major” items are indicated in bold italics.
- Check “none” for category only if there are no disease activity for the category
- For all study visits record the disease activity present within the 28 days prior to the visit.

There are 9 organ systems, plus an “Other” category for BVAS assessment. ***Major items are indicated in bold italics.***

1. General
   - Myalgia
   - Arthralgia / arthritis
   - Fever ≥38 °C
   - Weight loss ≥2 kg

2. Cutaneous
   - Infarct
   - Purpura
   - Ulcer
   - ***Gangrene***
   - Other skin vasculitis

3. Mucous membranes / eyes
   - Mouth ulcers
   - Genital ulcers
   - Adnexal inflammation
   - Significant proptosis
   - ***Scleritis / Episcleritis***
   - Conjunctivitis / Blepharitis / Keratitis
   - Blurred vision
   - Sudden visual loss
   - Uveitis
   - ***Retinal changes (vasculitis / thrombosis / exudate / haemorrhage)***

4. ENT
   - Bloody nasal discharge / crusts / ulcers / granulomata
   - Paranasal sinus involvement
   - Subglottic stenosis
   - Conductive hearing loss
   - ***Sensorineural hearing loss***

[redacted]

5. Chest
    - Wheeze
    - Nodules or cavities
    - Pleural effusion / pleurisy
    - Infiltrate
    - Endobronchial involvement
    - ***Massive haemoptysis / alveolar haemorrhage***
    - ***Respiratory failure***

6. Cardiovascular
    - Loss of pulses
    - Valvular heart disease
    - Pericarditis
    - Ischemic cardiac pain
    - Cardiomyopathy
    - Congestive cardiac failure

7. Abdominal
    - Peritonitis
    - Bloody diarrhea
    - ***Ischemic abdominal pain***

8. Renal
    - Hypertension
    - Proteinuria >1+ or >0.2 g/g creatinine
    - Hematuria ≥10 RBCs/hpf
    - Serum creatinine 125-249 µmol/L
    - Serum creatinine 250-499 µmol/L
    - Serum creatinine ≥500 µmol/L
    - ***Rise in serum creatinine >30% or fall in creatinine clearance >25%***

9. Nervous system
    - Headache
    - ***Meningitis***
    - Seizures (not hypertensive)
    - ***Cerebrovascular accident***
    - Organic confusion
    - ***Spinal cord lesion***
    - ***Cranial nerve palsy***
    - ***Sensory peripheral neuropathy***
    - ***Mononeuritis multiplex***

10. Other
    - ***RBC casts and/or glomerulonephritis***

#### 6.2 Disease Remission

BVAS scores will be used to assess disease remission at a respective timepoint.

[redacted]

- Remission at Week 26: BVAS of 0, and not taking glucocorticoids for treatment of AAV within 4-weeks of the Week 26 visit
- Sustained remission at Week 52: Remission at Week 26 without relapse to Week 52, AND remission at the Week 52 visit (BVAS of 0 and not taking glucocorticoids for treatment of AAV for 4 weeks prior to the Week 52 visit)

#### 6.3 Disease Relapse

A CEC Relapse AF must be completed for the Week 52 visit. On this form it must be documented whether the patient had any relapses between Week 26 and Week 52.

A relapse is defined as worsening of disease, after having previously achieved remission at week 26 (BVAS = 0 and having received no glucocorticoids for treatment of vasculitis for 4 weeks), that involves:

- one or more major item in the BVAS, or
- three or more minor items in the BVAS, or
- one or two minor items in the BVAS recorded at two consecutive study visits

***Major items are indicated in bold italics in Section 6.1***

#### 6.4 Vasculitis Damage Index

The Vasculitis Damage Index (VDI) is for recording organ damage that has occurred in patients since the onset of vasculitis.

Damage is defined as the presence of non-healing scars and does not give any indication of current disease activity. Damage items in the VDI are often the direct result of previous disease activity (captured in the BVAS). Damage is defined as having been present or currently present for at least 3 months. It is therefore possible for abnormalities to have occurred in the past, not be currently present, but to still count as damage.

Patients often have co-morbidity before they develop vasculitis, which must not be scored.

Record features of active disease using the BVAS, not the VDI.

New patients should usually have a VDI score of zero, unless:

a. They have had vasculitis for more than three months of onset of disease, and
b. The damage has developed or become worse since the onset of vasculitis.

The VDI item list can only deteriorate or be stable over time (damage is defined as irreversible in this scoring system). For each item in turn, record all features which have occurred since the onset of vasculitis, regardless of the cause.

[redacted]

For specific events, such as GI surgery, damage can be scored as positive if the procedure was undertaken at least three months prior to the assessment (and also must have occurred after the onset of the disease).

The same time frame is applied to all the damage items. If the patient is seen for the first time, and their vasculitis onset date is within three months of the assessment, then by definition, the patient cannot be recorded as having any damage. However, any features which are observed can be recorded as ascribable to damage after the arbitrary time of three months has elapsed.

##### 6.4.1 AAV Organ Systems in VDI

There are 11 organ systems in the VDI:

1. Musculoskeletal
   - Significant muscle atrophy or weakness
   - Deforming / erosive arthritis
   - Osteoporosis / vertebral collapse
   - Avascular necrosis
   - Osteomyelitis

2. Skin/Mucous Membranes
   - Alopecia
   - Cutaneous ulcers
   - Mouth ulcers

3. Ocular
   - Cataract
   - Retinal change
   - Optic atrophy
   - Visual impairment / diplopia
   - Blindness in one eye
   - Blindness in a second eye
   - Orbital wall destruction

4. Ear, Nose & Throat
   - Hearing loss
   - Nasal blockage / chronic discharge/crusting
   - Nasal bridge collapse / septal perforation
   - Chronic sinusitis / radiological damage
   - Subglottic stenosis (no surgery)
   - Subglottic stenosis (with surgery)

5. Pulmonary
   - Pulmonary hypertension
   - Pulmonary fibrosis
   - Pulmonary infarction
   - Pleural fibrosis
   - Chronic asthma
   - Chronic breathlessness

[redacted]

- Impaired lung function

6. Cardiovascular
   - Angina angioplasty
   - Myocardial infarction
   - Subsequent myocardial infarction
   - Cardiomyopathy
   - Valvular disease
   - Pericarditis ≥3 months or pericardectomy
   - Diastolic BP ≥95 or requiring antihypertensives

7. Peripheral Vascular Disease
   - Absent pulses in one limb
   - Second episode of absent pulses in one limb
   - Major vessel stenosis
   - Claudication >3 months
   - Minor tissue loss
   - Major tissue loss
   - Subsequent major tissue loss
   - Complicated venous thrombosis

8. Gastrointestinal
   - Gut infarction / resection
   - Mesenteric insufficiency / pancreatitis
   - Chronic peritonitis
   - Esophageal stricture / surgery

9. Renal
   - Estimated / measured GFR ≤50
   - Proteinuria **≥0.5** g/24 hours
   - End stage renal disease

10. Neuropsychiatric
    - Cognitive impairment
    - Major psychosis
    - Seizures
    - Cerebrovascular accident
    - 2nd cerebrovascular accident
    - Cranial nerve lesion
    - Peripheral neuropathy
    - Transverse myelitis

11. Other
    - Gonadal failure
    - Marrow failure
    - Diabetes
    - Chemical cystitis
    - Malignancy
    - Etc.

[redacted]

##### 6.4.2 Scoring of VDI

The VDI form was completed at the visits specified in the protocol Time and Events Table. In Protocol Section 12.4, instructions for completion, the organ systems with individual damage items, and the scoring of the VDI are provided.

The number of positive items is added for the total VDI score. Any previously scored items on the VDI must be carried forward to subsequent visits. The VDI score cannot decrease over time. The investigators did not need to calculate the VDI. This has been done in the EDC.

BVAS scoring for current disease activity can require assessment of the VDI profiles to distinguish active disease from cumulative organ damage occurring since the onset of disease. The VDI item list can only deteriorate or be stable over time (damage is defined as irreversible in this scoring system).

#### 6.5 Special Considerations/Conventions to Review Carefully PER PATIENT

- Low-dose steroid use and indication (see Glucocorticoid Rules/Conventions in section 5.2)

- Unscheduled BVAS assessments and ET visits:

    - If a scheduled visit is not available, Unscheduled and ET visits will be assigned to analysis visits using analysis visit windows based on the actual assessment date
    - The analysis window for Week 26 assessment is from Day 149 to Day 228 (inclusive).
    - The analysis window for Week 52 assessment is from Day 320 to Day 420 (inclusive).
    - The 4-week period prior to Week 26 is defined as 28 days prior to the actual Week 26 assessment date.
    - The 4-week period prior to Week 52 is defined as 28 days prior to the actual Week 52 assessment date.

- ET visit BVAS and/or VDI data will be adjudicated if applicable.

[redacted]

### 7. CEC Process Flow

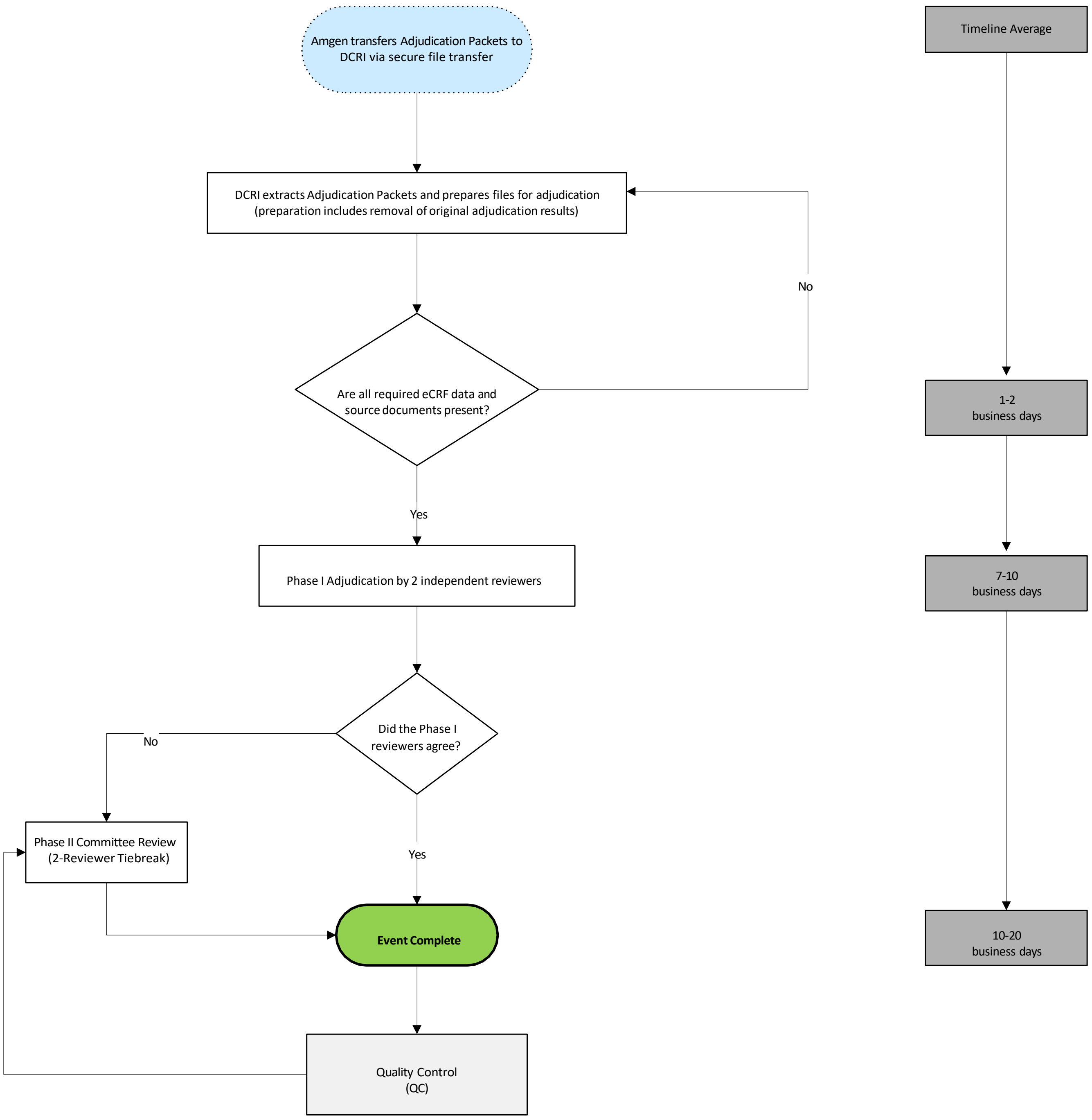

[redacted]
